# Global variation in cardiometabolic risk structures: A 48-country comparative Bayesian network analysis in 146,000 participants using WHO STEPS data

**DOI:** 10.64898/2026.05.15.26353288

**Authors:** Masih A. Babagoli, Michael J. Beller, Marco Scutari, Juan P. Gonzalez-Rivas, Jarvis C. Noronha, Andrea Medina, Natalia Sulbaran, Serina S. Cabrera, Aida Fallahzadeh, Sushruta Iruvanti, Ramfis Nieto-Martínez, Jeffrey I. Mechanick

## Abstract

**Background:** Cardiometabolic-based chronic disease (CMBCD) at an individual level results from complex interactions among a multi-tiered network of sociodemographic, behavioral, and metabolic factors. Though a consensus set of risk factors drives CMBCD, population context influences risk factor effects and interactions. To better understand this phenomenon, we investigated the multi-tiered networking of cardiometabolic variables across diverse populations using a comparative modelling approach.

**Methods and Findings:** Utilizing nationally representative cross-sectional data from 48 countries participating in the World Health Organization “STEPwise approach to noncommunicable disease risk factor surveillance” survey, we learned country-specific Bayesian networks including sociodemographic, behavioral, and cardiometabolic variables (adiposity, diabetes, hypertension, hyperlipidemia, and cardiovascular disease). By computing the structural Hamming distance between pairs of networks, we compared differences in network structures across regions and country income levels. We then used the learned networks to assess individual risk factor influences and interactions on cardiometabolic outcomes. Country-specific Bayesian networks varied in terms of the risk factors directly and indirectly associated with the cardiometabolic outcomes. Network structures differed significantly across regions (p = 0.023) but not across income levels (p = 0.91). These results were robust to an alternative learning algorithm, network comparison metric, and data imputation approach. Older age (60+ vs. 30-44 years old) was associated with a greater increase in probability of obesity in Europe and Central Asia (+80%) compared to other regions. Higher education was associated with increased probability of obesity (+53%), diabetes (+18%), and hypertension (+2%) in South Asia but decreased probability of obesity (−10%), diabetes (−32%), hypertension (−16%), and hyperlipidemia (−25%) in Middle East and North Africa. The interaction between age and sex in predicting obesity was significant in the highest proportion of countries in Europe and Central Asia compared to other regions. While this dataset provided standardized data across multiple countries to define cardiometabolic risk factors and drivers, there was limited data on certain health outcomes and uneven availability of data across regions.

**Conclusions:** These results revealed specific regional patterns of multi-tiered cardiometabolic risk structures, emphasizing the need for regionally tailored public health strategies rather than applying generalized consensus evidence-based models. Future research should explore the structural drivers of regional differences in inter-relationships of cardiometabolic risk factors, drivers, and disease.

## Introduction

Cardiometabolic-based chronic disease (CMBCD) imposes a growing global health burden, disproportionately affecting low- and middle-income countries.[1–3] As posited by several models, CMBCD is based on complex interactions of multiple tiers of sociodemographic, behavioral, and metabolic factors. Specifically, the CMBCD model incorporates the interaction of three primary drivers (genetics, environment, and behavior) and four secondary drivers (adiposity-, dysglycemia-, hypertension-, and lipid-based chronic disease) for certain cardiovascular diseases (CVDs; atherosclerosis, heart failure, and atrial fibrillation).[3] Importantly, these individual factors and their associated disparities are influenced by tertiary drivers such as social (socioeconomic status, education, housing, etc.), structural (policy and politics shaping individual context), and cultural determinants of health.[4, 5] Though a consensus set of risk factors is globally recognized in driving CMBCD, population differences in these drivers may impact how they interact to influence clinical cardiometabolic outcomes.

Previous epidemiological studies have reported the population attributable fraction (PAF) of CVD,[6–8] myocardial infarction,[9] and diabetes[10] associated with common cardiometabolic risk factors and compared those values across regions or country income levels. For example, Lin et al showed that while the mortality rate of diabetes attributable to high body mass index and overall dietary habits was higher than that attributable to other risk factors among countries of all income levels, the impact of other risk factors – including household air pollution and a diet high in processed meats – differed by country income level.[10] Yusuf et al showed in the INTERHEART study that nine risk factors together explained the majority of myocardial infarction risk in all regions and ages, though the PAF of myocardial infarction associated with each risk factor differed by region.[9] The PURE study, a global prospective cohort study that investigated the PAF of CVD associated with various risk factors, demonstrated that metabolic risk factors were the predominant CVD risk factors globally.[8] However, hypertension was associated with the highest PAF in low- and middle-income countries, while hyperlipidemia was associated with the highest PAF in high-income countries.[8]

The aforementioned studies have underscored that risk does not operate uniformly across settings, yet they predominantly focused on individual risk factors. Moreover, none have utilized a network-based approach to analyze the complex interactions among the multiple tiers of risk factors and compare these inter-relationships across multiple countries. Bayesian networks (BNs) can address this research gap. Specifically, BNs are probabilistic graphical models that represent variables (referred to as nodes) and the conditional dependencies between them (referred to as edges) as a directed acyclic graph.[11–13] The structure of these networks can be learned from data while also integrating prior knowledge in the form of imposed or disallowed edges. Analysis of BN structures can reveal inter-relationships among variables, and queries of the learned networks can demonstrate the relative impacts of one or more variables. As such, BNs have been used in multiple studies of CMBCD, albeit in the context of a single country.[14–22]

In this study, we employed BNs to analyze nationally representative cross-sectional data from 48 countries and investigate how multi-tiered networks of variables associated with CMBCD varied across settings. Specifically, we 1) learned country-specific BNs using sociodemographic, behavioral, and cardiometabolic variables, 2) compared the topology of BNs grouped by region and income levels, and 3) utilized the learned BNs to assess the interaction and impact of individual risk factors on clinical outcomes.

## Methods

### Data source

This study was a secondary analysis of the World Health Organization (WHO) “STEPwise approach to noncommunicable disease risk factor surveillance” (STEPS) survey.[23] STEPS is a standardized survey, developed by the WHO and carried out by individual country teams, that allows data collection of key behavioral and biological factors associated with non-communicable diseases for comparison across time and countries. The core survey comprises a questionnaire to capture basic sociodemographic information and medical history, as well as physical measurements and blood samples to ascertain adiposity, diabetes, hypertension, and hyperlipidemia status. The majority of country surveys employ a multi-stage complex sampling design to obtain nationally representative data.[23]

For this analysis, all country datasets from the last 15 years (2009-2024) were requested from the WHO NCD Microdata Repository [https://extranet.who.int/ncdsmicrodata/index.php/home]. For countries with multiple surveys, the most recent dataset was used. Country datasets were excluded from the analysis if: 1) any of the physical or biochemical measures used to define adiposity, diabetes, hypertension, and hyperlipidemia variables were greater than 40% missing, 2) the country dataset was not accessible through the WHO NCD Microdata Repository, or 3) if the dataset was subnational. See Supplementary Table 1 for a list of surveys included in this analysis. Inclusion criteria of individuals were: 1) age greater than or equal to 30 years and 2) not pregnant at the time of interview.

### Model variables

Model variables for inclusion in the BN were chosen based on the relevance to CMBCD and the availability of STEPS data across most country datasets. The following 14 variables were included: age, sex, urban/rural residence, education, household income, tobacco use, alcohol use, dietary fruit and vegetable intake, physical activity, adiposity, diabetes (type 1 and type 2 were not differentiated), hypertension, hyperlipidemia, and CVD (history of myocardial infarction or stroke). These variables were defined based on appropriate STEPS dataset variables, and differences across country surveys were harmonized. See Supplementary Table 2 for full definitions of model variables. In STEPS data collection, sex was recorded as binary “as observed”[23] by a data collector; this may not align with participant’s own identity and is a limitation of the data. Some variables, such as household income and alcohol use, were not available for all countries or had a high degree of missingness. Therefore, for each country dataset, a specific model variable was eliminated if there was greater than 40% missingness of that variable if it was one of the sociodemographic or behavioral variables. See Supplementary Table 3 for model variable inclusion for each country dataset.

### Data imputation

After excluding datasets and variables with missingness greater than aforementioned thresholds, the remaining missing values were imputed using a random forest algorithm[24] To optimize the out-of-bag error, a range of values was tested for the two parameters that correspond to the number of trees in each forest and the number of variables randomly sampled at each split. For each country, the parameters resulting in the lowest out-of-bag error were used to impute the variables. See Supplementary Tables 3 and 4 for the proportions of missing variables and the imputation error.

### Bayesian network structure learning

The 14 variables were organized into the following hierarchical order from highest to lowest tier: non-modifiable sociodemographic variables (age, sex, and urban/rural residence), modifiable sociodemographic variables (education and household income), behavioral variables (tobacco use, alcohol use, dietary fruit and vegetable intake, and physical activity), cardiometabolic drivers (adiposity, diabetes, hypertension, and hyperlipidemia), and CVD (history of CVD). Edge blacklists were defined to incorporate prior knowledge and improve the causal interpretation of the resulting networks. Notably, edges from lower-tiered variables to higher-tiered variables as well as edges among non-modifiable sociodemographic variables were disallowed. Additionally, edges among the cardiometabolic drivers were disallowed based on previous evidence regarding causal relationships among drivers.[3, 25]

For each country, a BN was learned using the hill-climbing algorithm with a bootstrap resampling approach modified to incorporate survey weights. Hill-climbing is a commonly used score-based graph structure learning algorithm that explores the space of possible directed acyclic graphs through sequential arc addition, deletion, and reversal steps in order to optimize fit according to a specified score.[26, 27] Subsequently, the learned edges are typically validated by bootstrap resampling of datasets, learning multiple networks, and constructing an averaged consensus network with edges that appear with a probability greater than a specific threshold.[28] To incorporate STEPS survey weights into this analysis, this methodology was modified based on an approach suggested by Gunawan et al.[29] Bootstrap resampling (n=1,000) of country datasets incorporated survey weights as probabilities to generate pseudo-representative samples for BN structure learning. Finally, a per-country averaged consensus network was derived with edges that were present in more than 50% of learned networks.

### Comparison of network topology

After learning a BN for each country, the network topologies were compared by calculating the structural Hamming distance (SHD) between each pair of networks. The SHD is the number of edge additions, deletions, and reversals needed to transform one network into another.[30] This metric has been previously used to compare learned BNs and quantify the difference between them.[31, 32] To analyze the factors associated with similarity in network topology, SHD values were compared based on whether the networks were from countries in the same or different World Bank region (East Asia and Pacific [EAP], Europe and Central Asia [ECA], Latin America and the Caribbean [LAC], Middle East and North Africa [MENA], South Asia [SA], or Sub-Saharan Africa [SSA]),[33] income level (low, lower-middle, upper-middle, or high),[33] and year of collection (pre-2017 or post-2017). For comparisons of SHD across regions, income levels, and years, unpaired two-sampled Wilcoxon tests were used, as the SHD was not normally distributed according to the Shapiro-Wilk test for normality.

### Bayesian network parameter learning

Given the learned BN structures, parameter learning was conducted using the corresponding dataset, incorporating the STEPS survey weights with implicit probabilistic expert system (I-PES) estimators suggested by Ballin and Vicard.[34, 35] Specifically, the I-PES estimator approximates the probability of each node in the BN integrating the BN dependence structure by conditioning on the parents of the node. This is done by calculating the conditional probabilities based on the country dataset frequencies weighted by the survey weights.

### Risk factor impact analysis

The learned BNs were then used for inference by using conditional probability queries to analyze the relative impact of sociodemographic and behavioral risk factors on the risk of CMBCD. Specifically, one of the nine sociodemographic and behavioral risk factors was instantiated to different values, and the relative change in the probability of the most advanced form of each cardiometabolic condition (obesity for the Adiposity node, diabetes for the Diabetes node, stage 2 hypertension for the Hypertension node, hyperlipidemia for the Hyperlipidemia node, and presence of CVD for the CVD node) was measured for each country. These queries were conducted using likelihood weighting, an approximate inference algorithm based on Monte Carlo sampling, by generating 10^7^ random samples. The median value of the relative change for each cardiometabolic condition was then calculated across all networks by region.

### Risk factor interaction analysis

To evaluate the interactions of risk factors, the per-country fitted BNs were used to generate a simulated dataset of the same size as the original dataset. For a given pair of risk factors and cardiometabolic condition, a “reduced” multinomial regression model was fitted with the risk factors as individual predictors and the cardiometabolic condition as the outcome. A “full” multinomial regression model was fitted adding an interaction term for the pair of risk factors as a predictor. To assess the overall significance of the interaction of those risk factors in predicting the outcome variable, we compared the “full” and “reduced” models using the likelihood ratio test; a p-value less than 0.05 was considered significant. This approach has been used in multiple similar analyses to assess interaction effects.[36, 37]

### Sensitivity analyses

Multiple sensitivity analyses were conducted to assess the robustness of differences in network topology across country datasets. First, BN structure learning was conducted using the tabu search algorithm instead of the hill-climbing algorithm. Tabu search is a modified version of hill-climbing and has been shown to have better performance in specific cases.[26, 38] Second, the degree of similarity between pairs of BNs was assessed by calculating the product-moment correlation between network pairs in addition to the SHD.[39] This approach was used in a previous analysis to compare BN structures.[32] Third, the incomplete observations in country datasets were imputed using multivariate imputation by chained equations (MICE) with polytomous regression instead of random forest imputation.[40] MICE is an imputation approach that has been shown to perform better than other methods for datasets that may have data missing not at random.[41]

### Statistical software

All analyses were performed using the R Statistical Software (version 4.2.2). BN structure and parameter learning were conducted using the *bnlearn* package.[42] Calculation of network centrality metrics and product-moment correlation were conducted using the *sna* package.[39]

### Ethical considerations

All STEPS country datasets received ethical approval from the appropriate local bodies, and informed consent was obtained from participants.[23] Our study team accessed de-identified licensed datasets through the WHO NCD Microdata Repository. Additional institutional review board approval was not required. This study was reported in accordance with the STROBE Statement for observational studies.

## Results

### Dataset characteristics

This study analyzed 48 STEPS country datasets spanning six regions and three income categories (Figure 1, Supplementary Table 1). The largest representation came from the EAP region (14 countries) and the lower-middle income group (20 countries). The median sample size of included datasets was 2,674 (range 365-7,295), with a total sample size of 146,086 across all countries.

**Figure 1.**
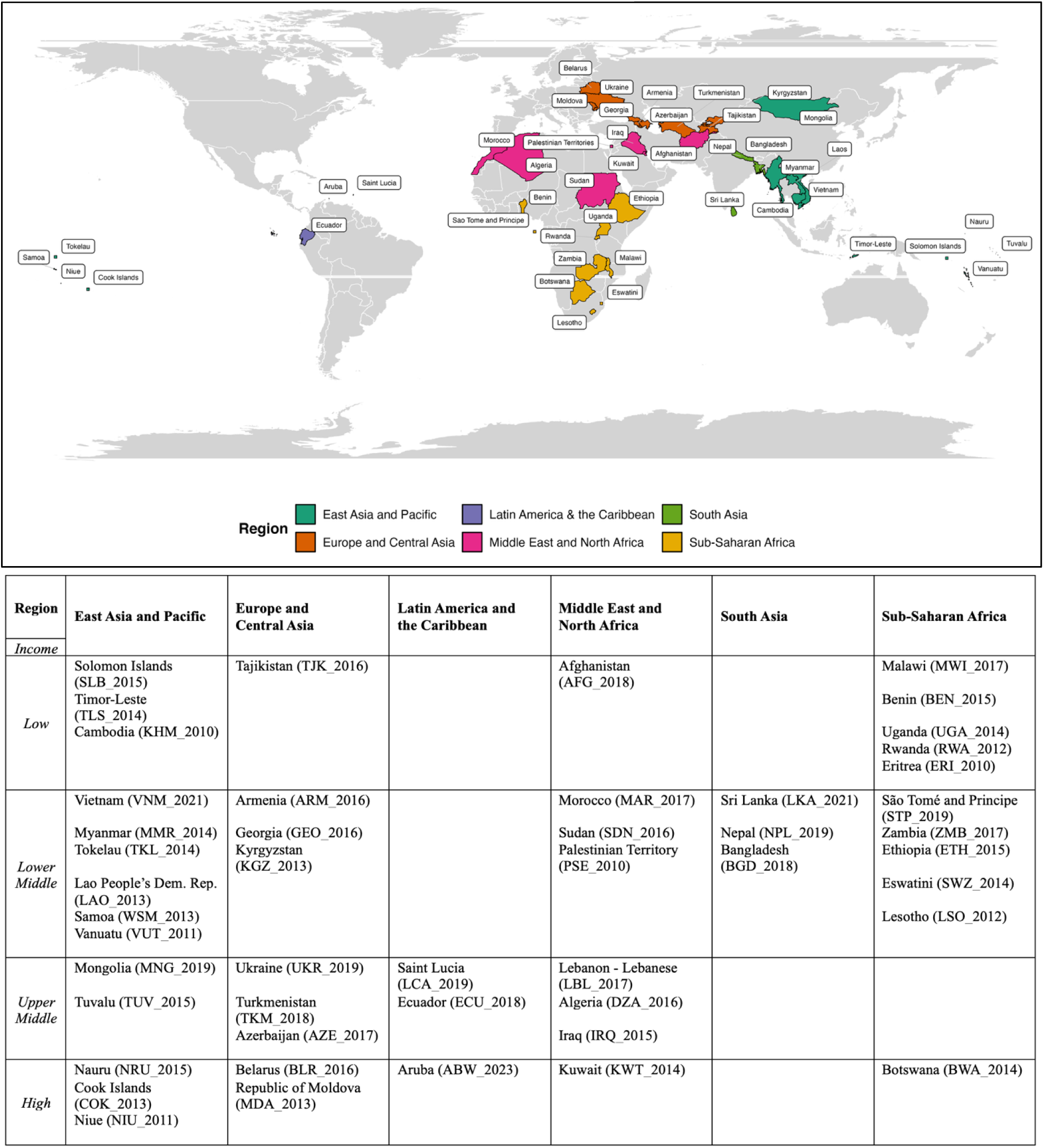
Country datasets included in analysis. Top Panel: Map of all country datasets included in this study. Bottom Panel: Breakdown of datasets by region and country income level. The dataset identification code in parentheses is used to identify the country dataset in other figures.

Fourteen model variables were defined and harmonized across datasets (Supplementary Figure 1, Supplementary Table 2). All 14 variables were available in 15 of 48 country datasets; the most frequently missing variable was urban/rural residence, absent in 25 country datasets (Supplementary Table 3). Across available variables, average data missingness per dataset was 0.9-13.6% (mean 4.1%). The mean out-of-bag error of the random forest imputation was 0.301 across all datasets (Supplementary Table 4).

### Bayesian network structure learning

BNs were learned for each of the 48 country datasets. These networks varied widely in the parent nodes of the cardiometabolic drivers and CVD (see Figure 2 for an example of network archetypes; see Supplementary Figure 2 for all networks). As examples, four of the 48 BNs are described. For Vietnam (VNM_2021), the parents of the cardiometabolic drivers of adiposity, diabetes, hypertension, and hyperlipidemia were primarily age, sex, and other drivers; additionally, tobacco use was a parent of adiposity. CVD was an isolated node (Figure 2a). In contrast, the network for Afghanistan (AFG_2018) had a more complex structure, with drivers linked both directly and indirectly to sociodemographic and behavioral variables. Adiposity was a child of physical activity and sex, which was a child of urban/rural residence and sex. CVD was a child of hypertension, which was a child of age and adiposity, and household income, which was a child of urban/rural residence and sex (Figure 2b). Similarly for Timor-Leste (TLS_2014), the direct parents of adiposity were age, physical activity, and alcohol use. However, adiposity was indirectly associated with sex, household income, and dietary fruit and vegetable intake through physical activity and alcohol use (Figure 2c). Finally, for Botswana (BWA_2014), age was a parent of diabetes, and diabetes was a parent of hyperlipidemia. Along with physical activity, hyperlipidemia was a parent of CVD. Meanwhile, household income and sex were parents of adiposity, which was a parent of hypertension (Figure 2d).

**Figure 2:**
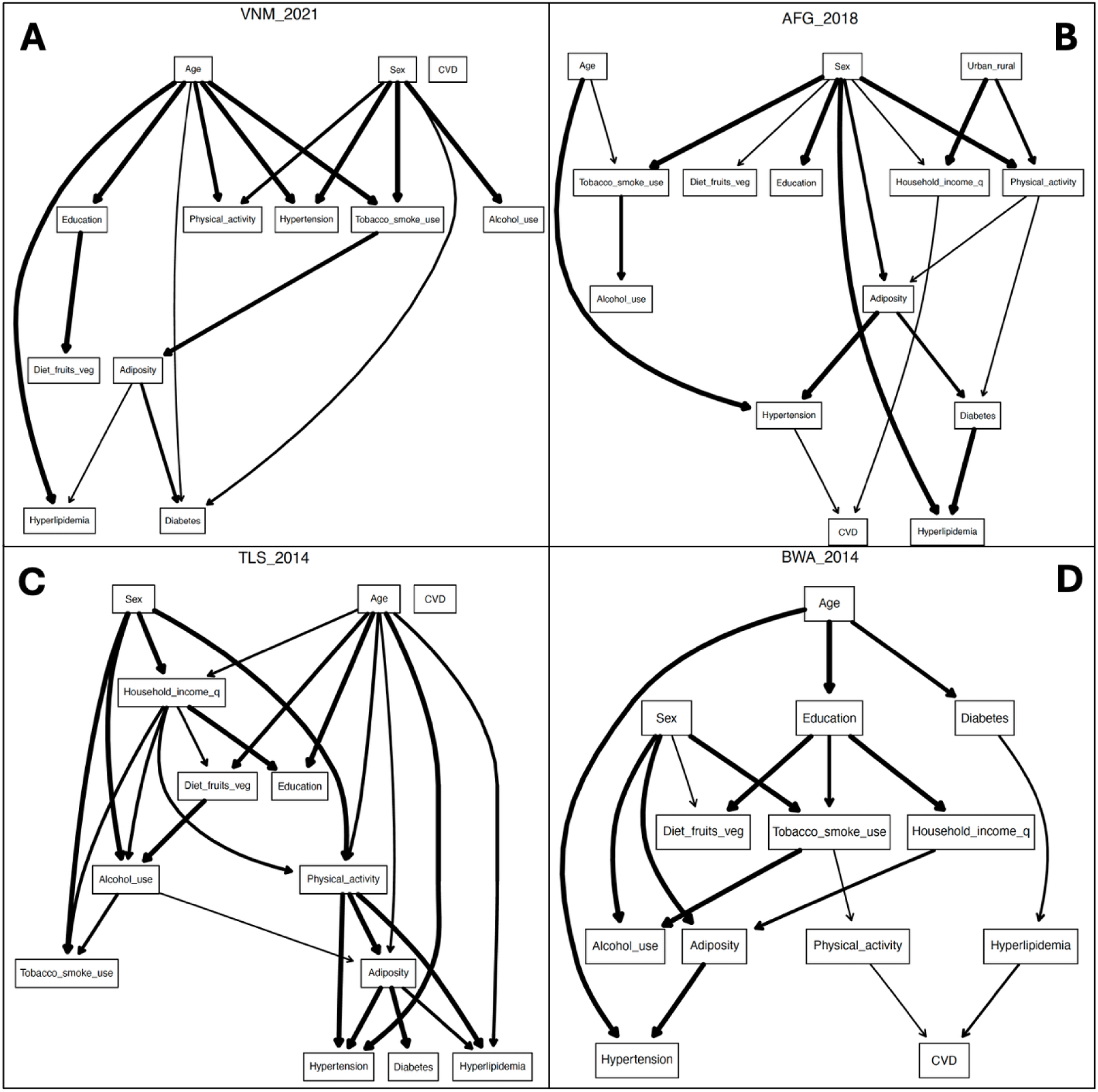
Bayesian networks learned for four different country datasets. Panel A - Vietnam (VNM_2021). Panel B - Afghanistan (AFG_2018). Panel C - Timor-Leste (TLS_2014). Panel D - Botswana (BWA_2014). Country-specific BNs varied widely in terms of the parent nodes of the cardiometabolic drivers and CVD. Edge thickness reflects strength of probabilistic relationships between two nodes, specifically based on the proportion of bootstrapped networks containing that edge. Edges present in more than 50% of bootstrapped networks were maintained for the averaged network for each country. Abbreviations: BN – Bayesian network; CVD – cardiovascular disease. See Supplementary Figure 2 for all 48 country networks.

### Comparison of network topology

The SHD was significantly lower between pairs of networks within the same region than between networks from different regions (Figure 3a; p-value 0.023). In contrast, the SHD did not differ for networks by country income level or year of dataset collection. These findings were robust in multiple sensitivity analyses using the tabu search for structure learning, comparing network topologies by the product-moment correlation, and imputing missing data with MICE approach (Supplementary Figures 4-6).

**Figure 3.**
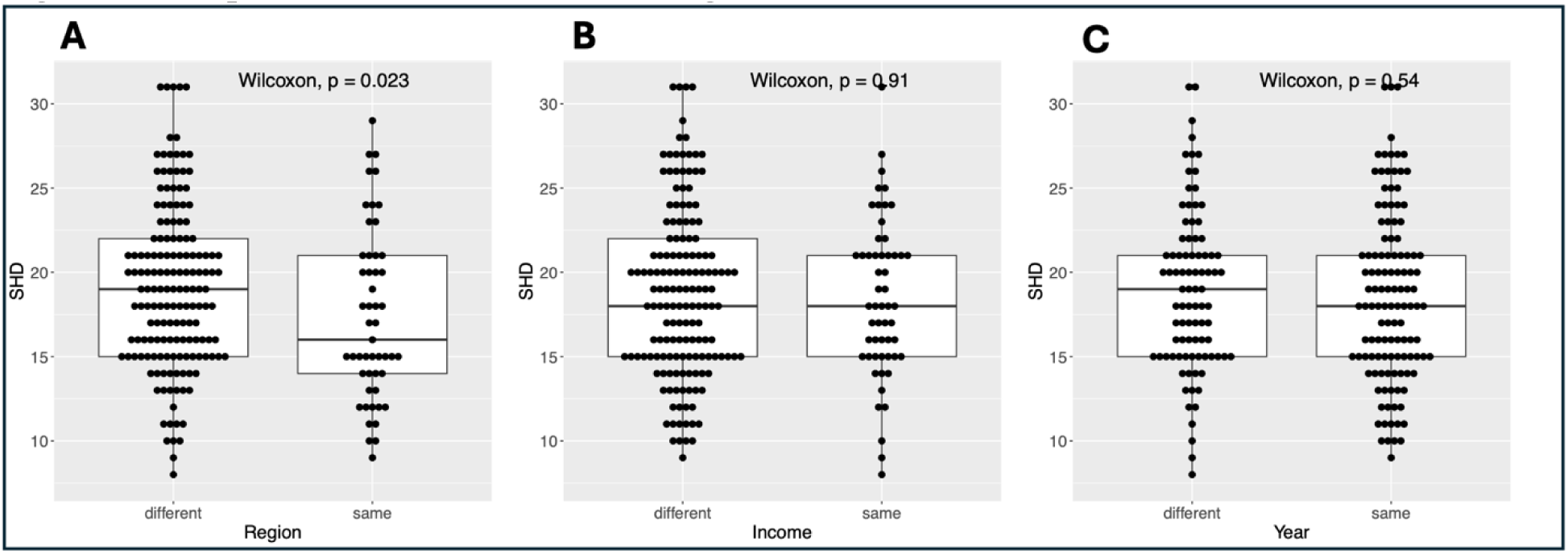
Comparison of structural Hamming distances. SHDs were compared for Bayesian networks: panel A – between same and different regions (p = 023); panel B – between same and different income levels (NS); and panel C – between same and different years of collection (NS). Data were imputed with Random Forest imputation, and network structure learned by hill-climbing algorithm. Each dot represents SHD for a pair of networks. Unpaired two-sampled Wilcoxon test used for comparison of means. Abbreviations: NS – not significant; SHD – structural Hamming distance.

### Conditional probability queries

Older age (60+ vs 30-44 years old) was associated with an increased probability of obesity in ECA (+83%) but showed marginal effect in other regions (Figure 4a). Across most regions, older age was associated with an increased probability of diabetes, hypertension, and hyperlipidemia. Finally, older age was associated with increased probability of CVD only in ECA (+182%). Compared to males, females had the largest relative increase in the probability of obesity compared to the other cardiometabolic outcomes across all regions (Figure 4b), with the largest relative increase in SSA (+175%) compared to other regions. Urban, compared to rural, residence was associated with an increased probability of obesity in SSA (198%) and EAP (+143%) and probability of hyperlipidemia in EAP (+205%) (Figure 4c).

**Figure 4.**
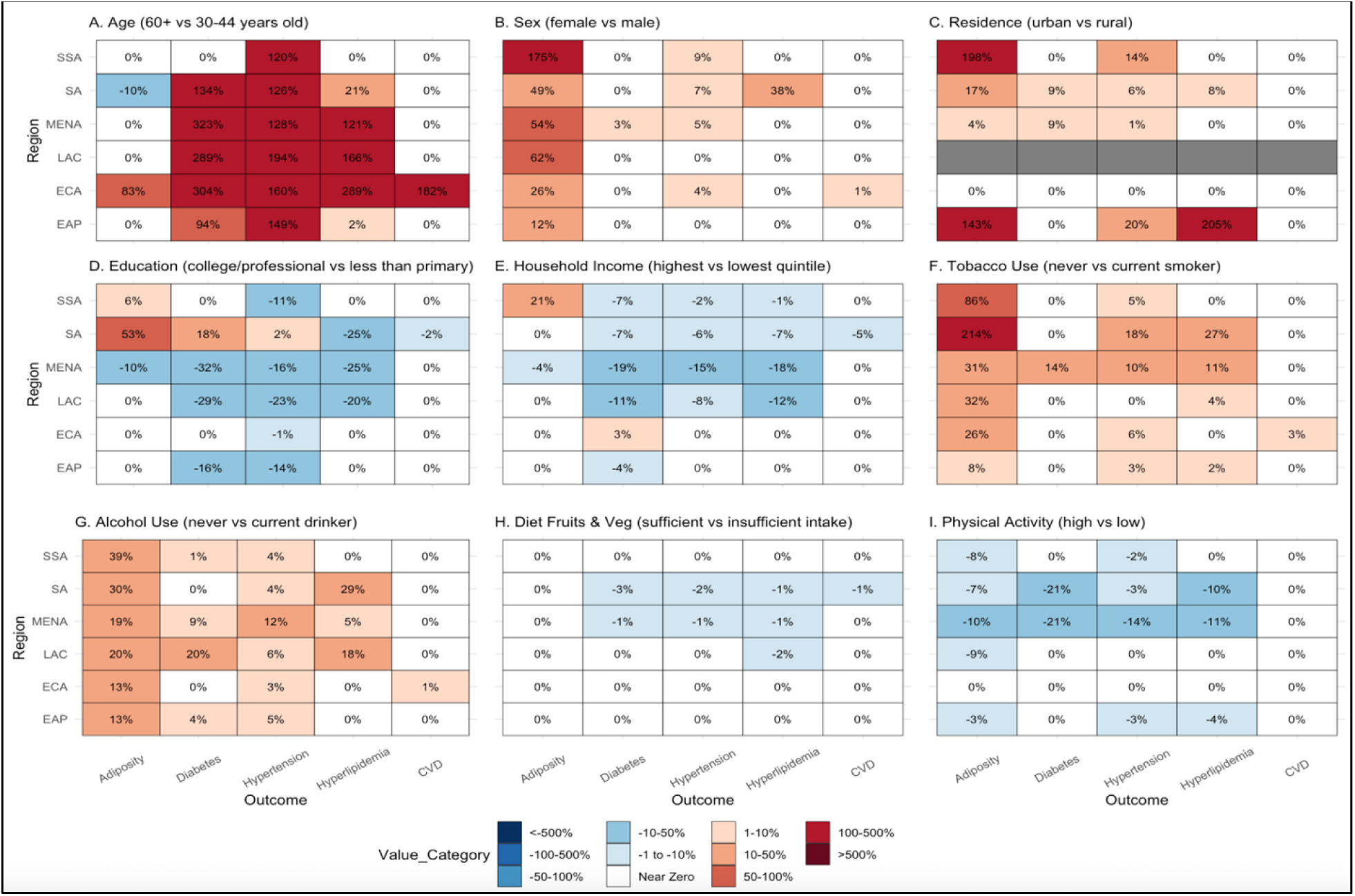
Conditional probability queries of single risk factors. Heat maps showing the relative impact of individual sociodemographic and behavioral risk factors on CMBCD. Percentages in each cell reflect the median relative change in probability of most advanced form of each cardiometabolic condition (obesity for Adiposity node, diabetes for Diabetes node, stage 2 hypertension for Hypertension node, hyperlipidemia for Hyperlipidemia node, and presence of CVD for CVD node) across Bayesian networks by region (left) and risk factor change (top). Values in headings indicate the starting and ending values of that specific risk factor that was instantiated. For instance, in panel A, the risk factor of age was changed from 30-44 to 60+ years old and the median relative change in each of the cardiometabolic outcomes across each region was calculated. Grayed out cells indicate variables that were not available for any country dataset in that region. Abbreviations: CMBCD – cardiometabolic-based chronic disease; EAP - East Asia and Pacific; ECA - Europe and Central Asia; LAC - Latin America & the Caribbean; MENA - Middle East and North Africa; SA - South Asia; SSA - Sub-Saharan Africa.

Higher education and household income were associated with increased probability of obesity in SSA (+6% and +21%, respectively), and higher education was associated with increased probability of obesity (+53%), diabetes (+18%), and hypertension (+2%) in SA (Figure 4d-e). In contrast, higher education and household income were associated with decreased probability of hyperlipidemia in SA (−25% and −7%, respectively) and decreased probability of obesity (−10% and −4%, respectively), diabetes (−32% and −19%, respectively), hypertension (−16% and −15%, respectively), and hyperlipidemia (−25% and −18%, respectively) in MENA (Figure 4d-e).

Never versus current tobacco use was associated with an increased probability of obesity in all regions (Figure 4f) with the largest relative increase in SA (+214%). Comparison of never to current alcohol use was associated with analogous findings, though the increase in probability of obesity was similar across all regions (Figure 4g). Sufficient fruit and vegetable intake was associated with minimal changes in the probability of all cardiometabolic outcomes (Figure 4h). Higher physical activity was associated with a decreased probability of cardiometabolic outcomes in specific combinations of outcome and region (Figure 4i). The greatest effect was observed in MENA, where higher physical activity was associated with a decreased probability of obesity (−10%), diabetes (−21%), hypertension (−14%), and hyperlipidemia (−11%).

### Risk factor interaction analysis

There was a significant interaction of age and sex in determining obesity in several countries (Figure 5). Comparing across regions, this interaction was present in the highest proportion of countries in ECA. Meanwhile, the interaction of age and sex in predicting hypertension was present in the highest proportion of countries in EAP. See Supplementary Figure 7 for the results of other risk factor pair interactions.

**Figure 5.**
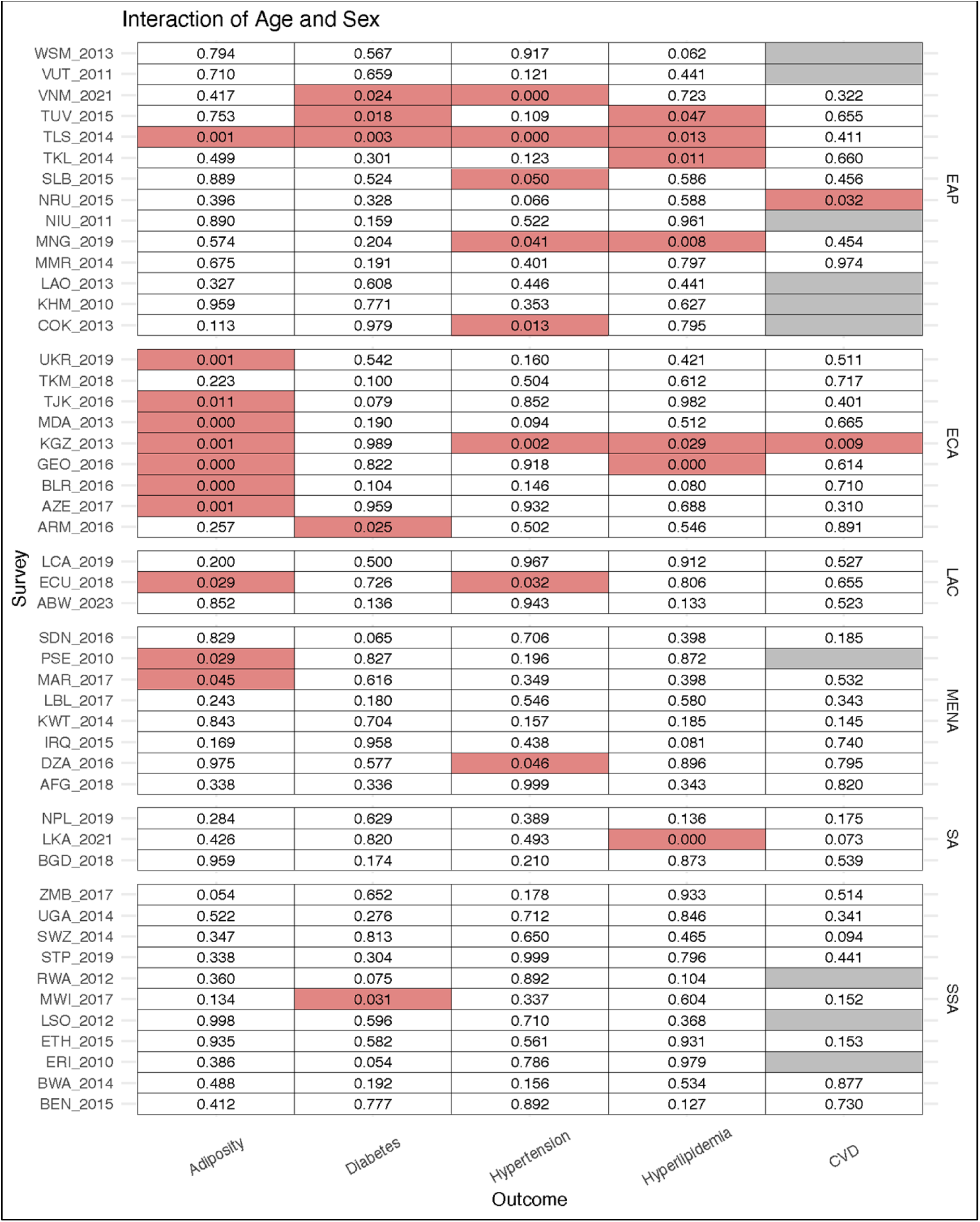
Interactions between Age and Sex in predicting cardiometabolic outcomes by country. Value in each cell reflects the p-value of the likelihood ratio test comparing the “full” and “reduced” multinomial regression models with Age and Sex as predictor variables and the cardiometabolic condition as the outcome variable. Models with a significant interaction between Age and Sex (p < 0.05) are shaded in red. Grayed out cells indicate variables that were not available for any country dataset in that region. Abbreviations: EAP - East Asia and Pacific; ECA - Europe and Central Asia; LAC - Latin America & the Caribbean; MENA - Middle East and North Africa; SA - South Asia; SSA - Sub-Saharan Africa. During the preparation of this work the author(s) used ChatGPT in order to assist with editing and troubleshooting code in R. After using this tool/service, the author(s) reviewed and edited the content as needed and take(s) full responsibility for the content of the published article.

## Discussion

To our knowledge, this is the first comparative network analysis of cardiometabolic risk factors, drivers, and disease across 48 countries using standardized nationally representative data. We found that 1) the learned country-specific BNs were more similar for countries in the same region; 2) the degree and direction of influence of certain risk factors on cardiometabolic outcomes differed by region; and 3) the interactions of specific pairs of risk factors on cardiometabolic outcomes also differed by region. Overall, these results reveal specific regional patterns of multi-tiered cardiometabolic risk structures, emphasizing the need for regionally tailored public health strategies.

Country-specific BNs of cardiometabolic risk factors and disease have been learned based on national datasets in the United States,[15, 43, 44] Spain,[16, 17, 19] and Korea[18] and utilized for various applications. Our analysis built on these prior studies by learning country-specific networks and then comparing the network structures. Network structures for each country varied significantly in terms of main risk factors associated with CMBCD outcomes, as well as the pathways through which modifiable and non-modifiable sociodemographic risk factors influenced clinical outcomes. Whether these learned networks reflect underlying differences in the causal mechanisms by which CMBCD develops in each country depends on certain assumptions. We approximated these assumptions by including all relevant variables available for each country survey while disallowing illogical directed edges and specific directed edges among cardiometabolic outcomes not supported by the medical literature. Even if the country-specific BNs learned from the data do not represent causal mechanisms, the network edges do reflect the strongest statistical associations between the variables of interest and, thus, patterns in the country-specific epidemiology of CMBCD. Therefore, our finding that the BNs of countries in the same region were more structurally similar than those of countries in different regions is remarkable and suggests specific regional effects interconnecting sociodemographic, behavioral, and cardiometabolic variables in country-specific networks.

Beyond differences in the overall network structure, we also demonstrated regional differences in the impact of risk factors and distinct interactions between pairs of risk factors on cardiometabolic outcomes. Education and/or household income were inversely associated with the probability of obesity in SA and SSA though not in other regions. This inverse association has been previously reported in countries of lower socioeconomic development, which characterizes many SA and SSA countries.[45–47]

Furthermore, we found a significant interaction between age and sex in determining the probability of obesity in the highest proportion of countries in ECA compared to other regions, suggesting that the impact of age on obesity differs by sex in that region. In Global Burden of Disease analyses of ECA, males had higher rates of obesity than females prior to 40 years old, but females had higher rates after that age.[48] These differential effects by sex in a particular region highlight an example of nuanced regional patterns in CMBCD epidemiology.

The underlying explanation for this regional effect is likely related to contextual differences in each part of the world – “hidden variables” that shape the network such as demographic distribution, cultural factors, and structural determinants (e.g. healthcare access, food-related policies, and environmental factors). Regardless of the explanation, the regional differences in the interplay of variables underlying CMBCD have implications in strategies to address the growing disease burden. Seminal studies have demonstrated that a consensus set of risk factors account for a large portion of conditions such as myocardial infarction and CVD, amounting to generic prevention principles worldwide.[8, 9] However, our findings indicate that prevention efforts could be strengthened by accounting for region-specific risk factor effects and interactions, allowing interventions to more effectively target high-risk groups and locally relevant drivers of CMBCD. This process is a critical aspect of “transculturalization” – the adaptation of clinical practice guidelines and other evidence-based frameworks from one setting to another in a manner that respects cultural and environmental differences.[49, 50]

On the other hand, certain patterns were present in all or most regions. Compared to all other risk factors, age had the most significant influence on probability of diabetes, hypertension, and hyperlipidemia in most regions. Meanwhile, never smokers and never drinkers had a higher probability of obesity than current smokers and drinkers, respectively, in all regions. This association between tobacco use and adiposity has been supported in some studies[51, 52] and refuted in others.[53] While the mechanism of this association is debated, neurobiological changes in weight regulation with smoking are often cited.[54] The association between alcohol use and adiposity has not been fully established, with some evidence suggesting that the relationship depends on the quantity of alcohol use.[55] Diet had minimal impact on all cardiometabolic outcomes in all regions. Given that diet is a well-established determinant of CMBCD, this finding is likely a limitation of our definition of diet solely based on fruit and vegetable intake. Alternatively, cardiometabolic outcomes may not be strongly linked with concurrent dietary patterns, which is a limitation of cross-sectional surveys.

### Strengths and Limitations

Our study has several strengths. To our knowledge, this study is the first to employ a country-specific network approach to analyze the multi-tiered drivers of CMBCD and compare results across a large set of countries. BNs uniquely allow modeling of the interdependencies among multi-tiered variables and the indirect effects of proximal risk factors on downstream health outcomes, an advantage that has been previously illustrated in the context of CMBCD.[44] Subsequent comparison of network topology extends conventional analyses to understand how the inter-relationships among variables differ across countries. Furthermore, this study asserts a notable methodological improvement compared to prior studies that utilized BNs to analyze CMBCD using large datasets collected via complex sampling designs. While such BN analyses disregarded the accompanying survey weights, we integrate validated methods into both the structure and parameter learning stages to incorporate STEPS survey weights and ensure more accurate estimation. Finally, in contrast to prior multinational studies that use pooled dataset to study cardiovascular risk factors, our approach to separately learn networks based on each country dataset provides nationally representative country-specific analyses. These learned country-specific BNs can be used by researchers and policymakers to conduct theoretical experiments and simulate the impacts of interventions.

In terms of limitations, while the STEPS dataset provided standardized data across multiple countries and many variables to broadly define cardiometabolic drivers, there was limited data on certain outcomes. Diabetes was not differentiated between type 1 and type 2. The questionnaires captured CVD as any history of heart attack or stroke. Other CVD manifestations were not included, and the lack of imaging precluded identification of subclinical disease. CVD history self-report is likely less accurate in low-resource settings with limited access to care. Additionally, the use of cross-sectional, rather than longitudinal, data may have underestimated the impact of long-term behavioral factors such as diet and physical activity. Though these limitations of the STEPS dataset are notable, they represent an expected tradeoff to the considerable scope and crucial cross-country standardization of the dataset. Finally, the STEPS data was not available for all countries, a limitation more pronounced in specific regions and income levels. Acknowledging that this may affect the averaged results, we included all regions and income levels with data to provide as global of an analysis as possible.

There were important limitations of the BN methodology. Structure-learning algorithms prioritize the strongest relationships to produce the best-fitting model. Consequently, weaker associations may be excluded from the learned structure and, thus, will not be reflected in conditional probability queries using the learned network. This likely explains the null effect of some risk factors on cardiometabolic outcomes present in our conditional probability queries. Finally, BNs were originally conceived as a generative model designed to simulate scenarios. Thus, there is limited literature on validated approaches to conduct hypothesis testing using the results of model queries. Our study utilizes conditional probability queries to illustrate the general trends on the impact of risk factors but not to make precise estimates of metrics such as PAF or odds ratios.

In this comparative network analysis of cardiometabolic risk factors, drivers, and disease across 48 countries, we found regional patterns of multi-tiered cardiometabolic risk structures and differential effects and interactions of specific risk factors. This finding emphasizes the need for regionally tailored public health strategies to effectively target high-risk groups.

## Data Availability

No data was generated by this study. The following existing data sources were used: STEPwise approach to noncommunicable disease risk factor surveillance (STEPS) from WHO NCD Microdata Repository available via https://extranet.who.int/ncdsmicrodata/index.php/home.

